# Evaluation of the disease-modifying effects of Aureobasidium pullulans AFO-202 strain produced Beta-Glucan in Parkinson’s disease – Results of a pilot clinical study

**DOI:** 10.1101/2023.04.14.23288571

**Authors:** Chockanathan Vetrievel, Allimuthu Nithyanandam, Subramaniam Srinivasan, Sudhakar S Bharatidasan, Vidyasagar Devaprasad Dedeepiya, Nobunao Ikewaki, Masaru Iwasaki, Rajappa Senthilkumar, Senthilkumar Preethy, Samuel JK Abraham

**Affiliations:** BeWell Hospitals, Chennai, India; Apollo First Med Hospital, Chennai, India; Mary-Yoshio Translational Hexagon (MYTH), Nichi-In Centre for Regenerative Medicine (NCRM), Chennai, India; Department of Anesthesia, Thunder Bay Regional Health Sciences Centre, Thunder Bay, Ontario, Canada; Dept. of Medical Life Science, Kyushu University of Health and Welfare, Japan; Institute of Immunology, Junsei Educational Institute, Nobeoka, Miyazaki, Japan; Centre for Advancing Clinical Research (CACR), University of Yamanashi - School of Medicine, Chuo, Japan; Fujio-Eiji Academic Terrain (FEAT), Nichi-In Centre for Regenerative Medicine (NCRM), Chennai, India; Antony- Xavier Interdisciplinary Scholastics (AXIS), GN Corporation Co. Ltd., Kofu, Japan; Sophy Inc., Nyodogawa, Kochi, Japan

**Author notes:** **Corresponding author information:** Dr. Samuel JK Abraham, II Department of Surgery & CACR, University of Yamanashi, Faculty of Medicine, Address for correspondence: 3-8, Wakamatsu, Kofu, 400-0866, Yamanashi, Japan, Email id-; Alternate email id. **<u>Availability of data and material</u>** <u>All data generated or analysed during this study are included in the article itself.</u>. **<u>Funding</u>** No external funding was received for the study. **<u>Competing interests</u>** Author Samuel Abraham is a shareholder in GN Corporation, Japan which holds shares of Sophy Inc., Japan., the manufacturers of novel beta glucans using different strains of Aureobasidium pullulans; a board member in both the companies and also an applicant to several patents of relevance to these beta glucans. **<u>Ethics Approval:</u>** The study was an open label, prospective, non-randomised, single arm pilot clinical study which was registered in India’s clinical trial registry CTRI (ref no: CTRI/2022/05/042498) and conducted following all prevailing national guidelines. Subjects provided written consent according to the Declaration of Helsinki.

**Keywords:** Parkinson’s disease (PD), Alpha-synuclein, Beta-glucans, UPDRS, Constipation, MRI

## Abstract

The aetiology of Parkinson’s disease (PD) has been linked to the aggregation and spread of misfolded alpha-synuclein via the gut-brain axis. We previously reported the effects of a biological response modifier, beta-glucan, produced by the AFO-202 strain of *Aureobasidium Pullulans*, which improves clinical symptoms and controls gut *Enterobacteriaceae* associated with curli and amyloid-alpha-synuclein production. In this study, we report the effects of beta-glucan on PD. Eight patients with PD were recruited, five of whom completed the study. Each participant was administered 3 g of AFO-202 B-glucan orally daily for 90 days in addition to their regular prescription drugs. Pre- and post-study comparison revealed that the mean UPDRS decreased from 43.25 ± 13.75 at baseline to 40 ± 13.65 post intervention. Improvements in cognition, walking and balance, postural stability, and constipation scales were observed. The mean constipation severity score decreased from 3 ± 1.73 to 1.75 ± 0.43 post intervention. The serum creatinine kinase levels decreased and the blood glucose and lipid levels normalised. The MRI Parkinson’s index (MRPI) improved in one patient. This safe AFO-202 B-glucan produced beneficial disease-modifying improvements in the UPDRS and MRI that were clinically significant in the short timeframe of 90 days. Further validation in larger, longer-term clinical trials will help confirm the use of beta-glucan as a potential adjuvant treatment for PD which may pave way for future evaluations of these beta-glucans in other synculeinopathies as well Lewy-body related pathogenesis.

## Introduction

Parkinson’s disease (PD) is a prevalent neurodegenerative condition characterised by bradykinesia, rest tremors, rigidity, postural instability, and a variety of other subtle motor symptoms and non-motor abnormalities [1,2]. The loss of dopaminergic neurons in the substantia nigra pars compacta is considered a key factor in motor problems [1,3]. Most current pharmacological PD therapy strategies focus on regaining striatal dopaminergic tone. Levodopa, a precursor of dopamine, was the first drug developed to treat PD in the 1960s and remains the most efficient treatment for the disease. Other dopaminergic medications have been employed, such as dopamine metabolism inhibitors and dopamine receptor agonists; however, these are typically less well tolerated and less efficient [1]. Owing to the distribution of dopamine in extra-striatal areas, heterogeneity in its absorption, and transit across the blood-brain barrier, existing therapies are associated with substantial side effects [1]. Therefore, developing disease-modifying therapies (DMT) is necessary to reduce these side effects.

The presence of aberrant α-synuclein aggregates is a major pathological sign that is now being increasingly associated with Parkinson’s disease (PD). α-Synuclein plays a central pathogenic role through vesicular transport disruption, alterations in the lysosome-autophagy system, mitochondrial dysfunction, and oxidative stress. It has been suggested that pathogenic forms of α-synuclein may behave similarly to prion diseases, allowing the pathology to propagate among cells [4, 5]. Any DMT that is being developed should be able to address the α-synuclein aggregation as well.

A biological response modifier beta-glucan from the AFO-202 strain of a black yeast called *Aureobasidium pullulans* has been reported to improve the behaviour and sleep pattern with increase in melatonin and α-synuclein levels in a clinical study done in children with autism. In these study participants, whole-genome metagenome (WGM) sequencing of stool samples revealed that among the relevant species, the prevalence of Enterobacteriaceae that produce curli, which causes α-synuclein misfolding and aggregation, was greatly decreased. Gut dysbiosis is also regulated by an increase in beneficial bacteria, such as *Faecalibacterium prausnitzii* and *Prevotella copri*, and a decrease in other harmful bacteria, such as *Escherichia coli, Akkermansia muciniphila, Blautia* spp., *Coprobacillus* sp., and *Clostridium bolteae*. Based on these findings, we studied the effects of AFO-202-glucan on patients with PD.

## Methods

This open-label, prospective, non-randomised, single-arm pilot clinical study was registered in India’s Clinical Trial Registry (ref no: CTRI/2022/05/042498) and conducted following all prevailing national guidelines. All participants provided written informed consent in accordance with the Declaration of Helsinki.

### Inclusion and exclusion criteria

Eligibility criteria included adults aged ≥18 years, diagnosed with Parkinson’s Disease that meet the clinical diagnostic criteria of the brain bank of the Parkinson’s Disease Association of the United Kingdom. Subjects were instructed to follow the same stable regimen of PD medications for a minimum of three months prior to enrolment and were willing to not make major changes to their standard treatment regimen until the end of the treatment period. Patients were excluded if they were in an advanced stage of PD or had a history of allergic reactions to the key constituents of the investigational product. Subjects with difficulty swallowing or any condition that made oral medication difficult or impossible were also excluded.

We excluded subjects who had undergone major surgical procedures within four weeks prior to randomisation, who were on antidepressants or antipsychotics, or presented with psychiatric conditions that would interfere with the parameters of the clinical study, subjects with a known history of clinically significant endocrine, gastrointestinal, cardiovascular, haematological, hepatic, immunological, renal, respiratory, or genitourinary abnormalities or diseases, except those that were considered aetiology or comorbid to the study indication, pregnant, lactating, or planning to become pregnant, and subjects who were participating or had participated in a clinical trial within 90 days of the start of the present trial.

### Intervention

All participants were asked to consume three sachets of AFO-202 beta-glucan powder (each sachet containing 1 g of 1,3-1,6 beta-glucan from *A. pullulans* AFO-202 strain) (commercial name: Nichi BRITE); one sachet was consumed 30 min after a meal, once in the morning, once in the afternoon, and once in the evening.

### Assessments

The following assessments were performed at baseline and end of the 90 days of intervention:

1. Unified Parkinson’s Disease Rating Scale (UPDRS)
2. Magnetic Resonance Imaging (MRI) of brain
3. Constipation Severity Score
4. Serum Creatinine Kinase
5. Fasting, Post-prandial blood sugar, HbA1c, Lipid levels in blood

### Safety Monitoring

The frequency and severity of adverse events and any clinically abnormal safety parameters were monitored via telephone conversations during the study and on the 90^th^ day.

### Target population for analysis

The intention-to-treat (ITT) analysis included all participants enrolled in the study. Per-protocol analysis (PPS) was performed on participants who completed the entire study.

### Method of analysis

All data were analysed using Excel (Microsoft Office Excel software®). Paired Student’s t-tests were also performed using this package, and p-values <0.05 were considered statistically significant.

## Results

Eight patients who met the inclusion criteria were included in the study and ITT analysis. During the study, three subjects were lost to follow-up, as one of them developed loose stools which were controlled with medication, and the other three developed difficulties in accessing the hospital for assessment because they were totally dependent on their caretakers.

The age and other demographics of the participants are shown in Table 1.

**Table 1.** Demographics of participants who completed the study

| Subject | Gender | Age (years) | Height (cm) | Weight (kgs) | Pulse rate | Respiratory rate (breaths per minute) | Year of diagnosis |
| --- | --- | --- | --- | --- | --- | --- | --- |
| 1 | M | 71-75 | 155 | 39 | 98 | 16 | 2019 |
| 2 | M | 8-85 | 167 | 68.8 | 72 | 17 | 2018 |
| 3 | M | 51-55 | 165 | 72.4 | 74 | 17 | 2020 |
| 4 | M | 86-90 | 155 | 51.5 | 118 | 16 | 1992 |
| 5 | M | 65-70 | 159 | 49.7 | 76 | 16 | 2020 |

The mean UPDRS score at baseline was 43.25 ± 13.75 which decreased to 40 ± 13.65 (p value = 0.5) after 90 days (Figure 1). Improvements were observed in cognition, walking and balance, postural stability, and constipation scales. The mean constipation severity score decreased from 3 ± 1.73 to 1.75 ± 0.43 post intervention (p-value =0.2) (Figure 2). The serum creatinine kinase level decreased from 189.19 ± 99.99 U/L to 125.52 ± 70.90 U/L. The mean total cholesterol decreased from 190.67 ± 31.65 mg/dL to 160.16 ± 28.69 mg/dL. Mean triglyceride levels at baseline decreased from 187.50 ± 218.81 to 112 ± 59.66 mg/dL. The mean LDL cholesterol decreased from 102.67 ± 27.93 to 95.83 ± 21.67 mg/dL. The mean VLDL decreased from 37.67 ± 43.67 to 22.5 ± 11.95 mg/dL (Figure 3). The mean fasting blood glucose decreased from 114.83 ± 35.40 to 88 ± 15.12 mg/dL. The mean postprandial blood glucose decreased from 203.33 ± 112.28 to 139.66 ± 42.92. The mean Hba1c decreased from 7.10 ± 1.55 to 6.68 ± 1.04 after 90 days (Figure 4). The ESR decreased from 17.33 3.68 to 16.50 1.55.

**Figure 1:**
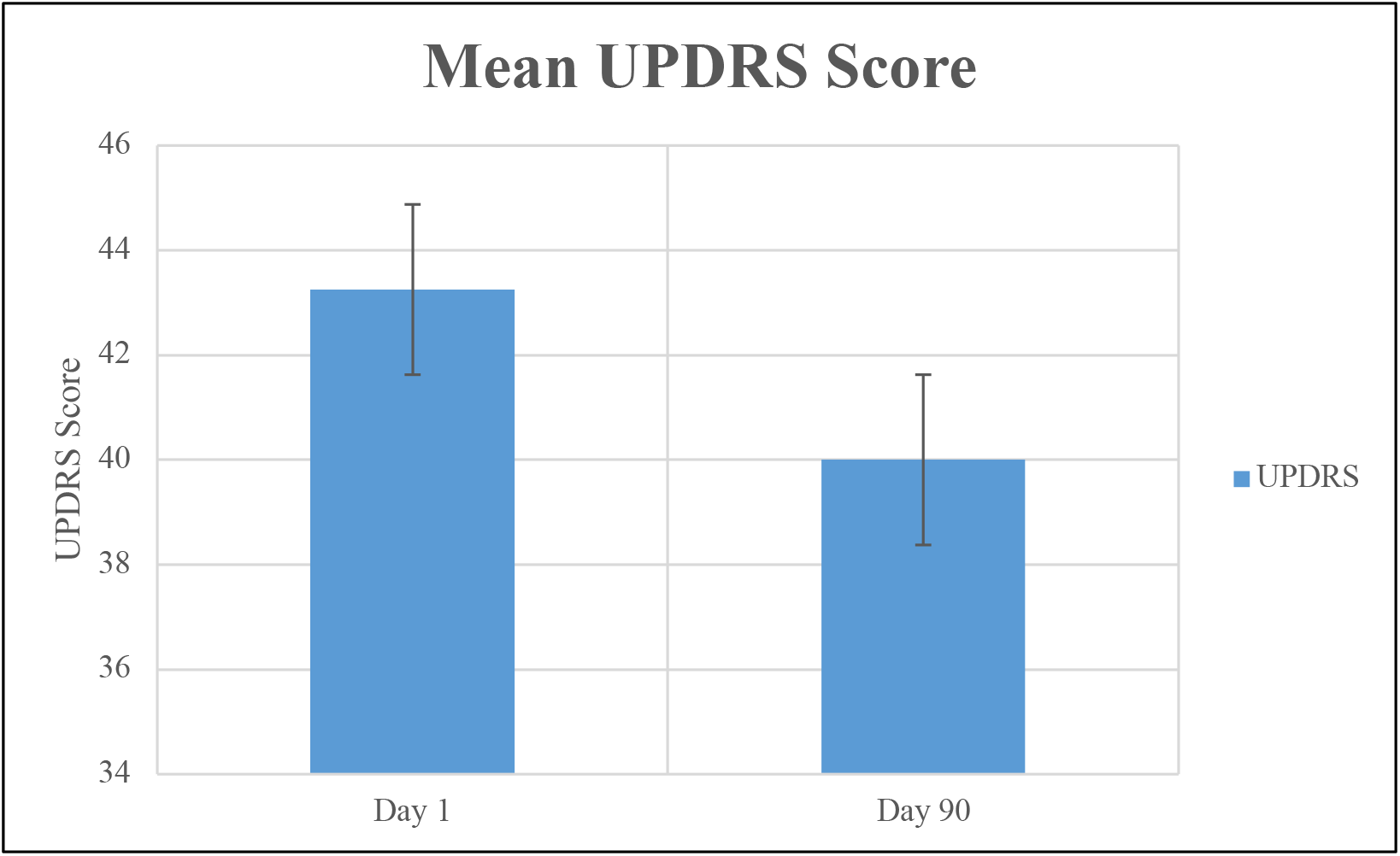
Mean Unified Parkinson’s Disease Rating Scale (UPDRS) at baseline and post intervention

**Figure 2:**
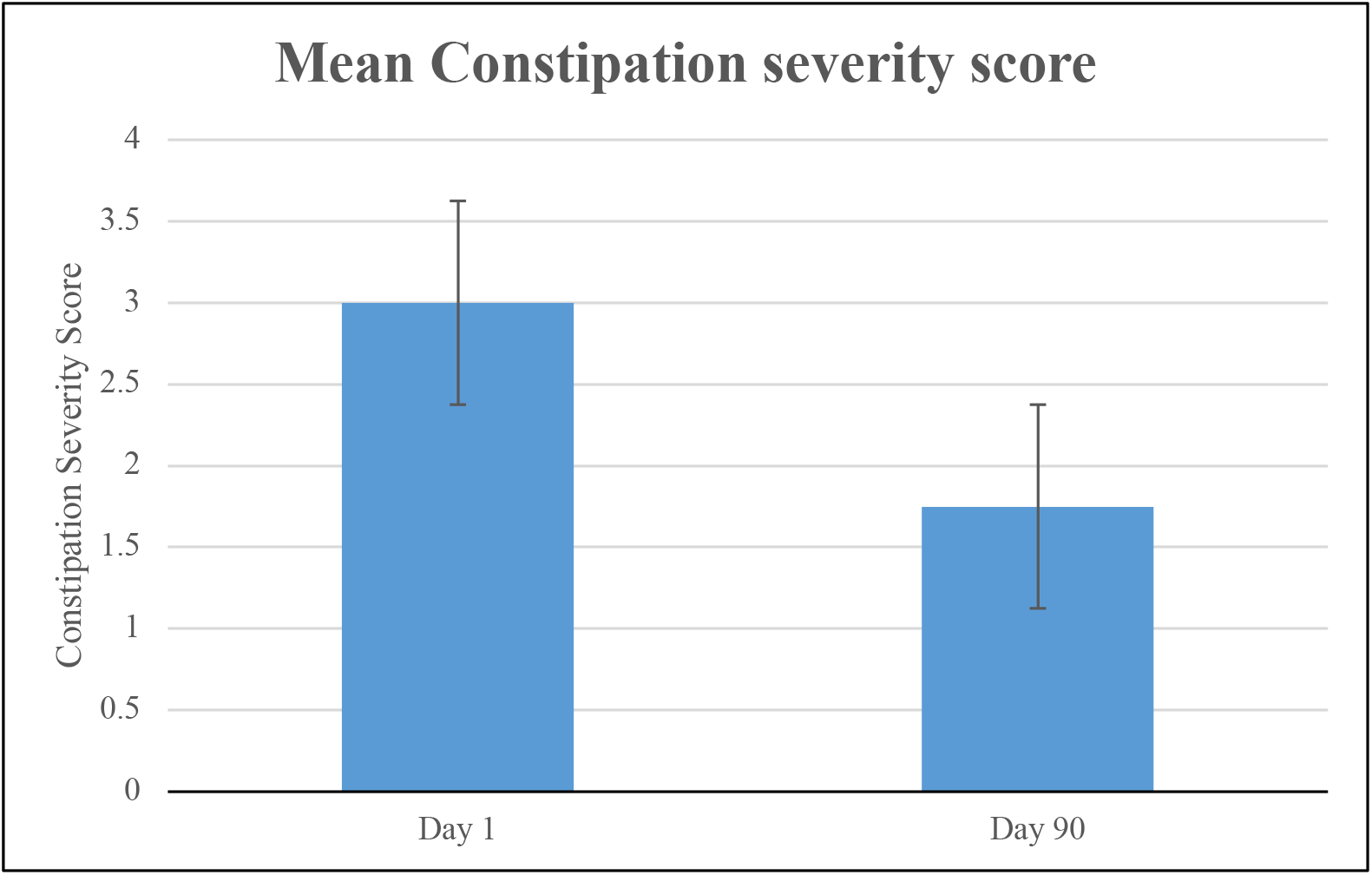
Mean constipation severity score, which decreased 90 days post consumption of AFO-202 beta-glucan

**Figure 3:**
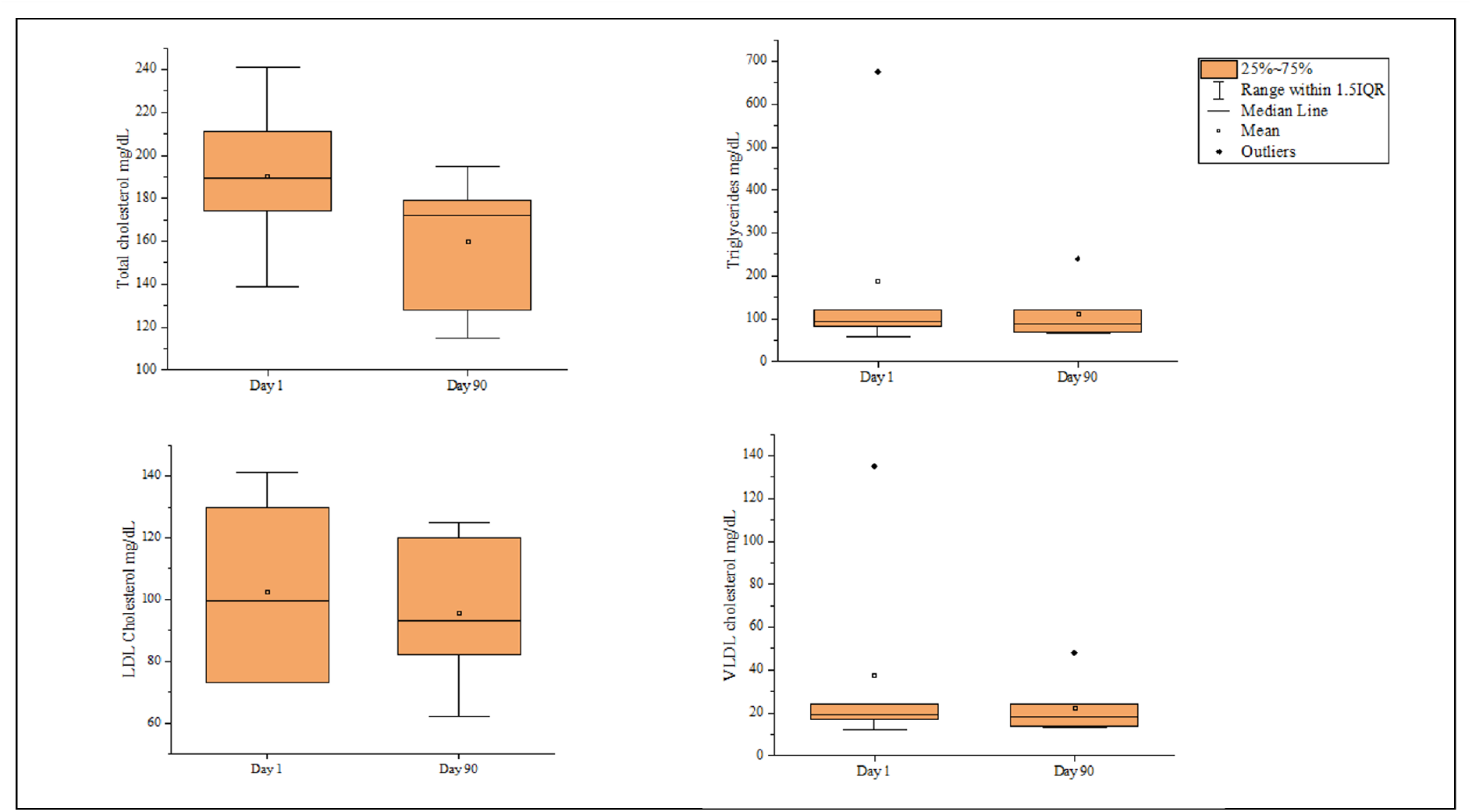
Values of total cholesterol, triglycerides, LDL cholesterol, and VLDL cholesterol pre and post consumption of AFO-202 beta-glucan

**Figure 4:**
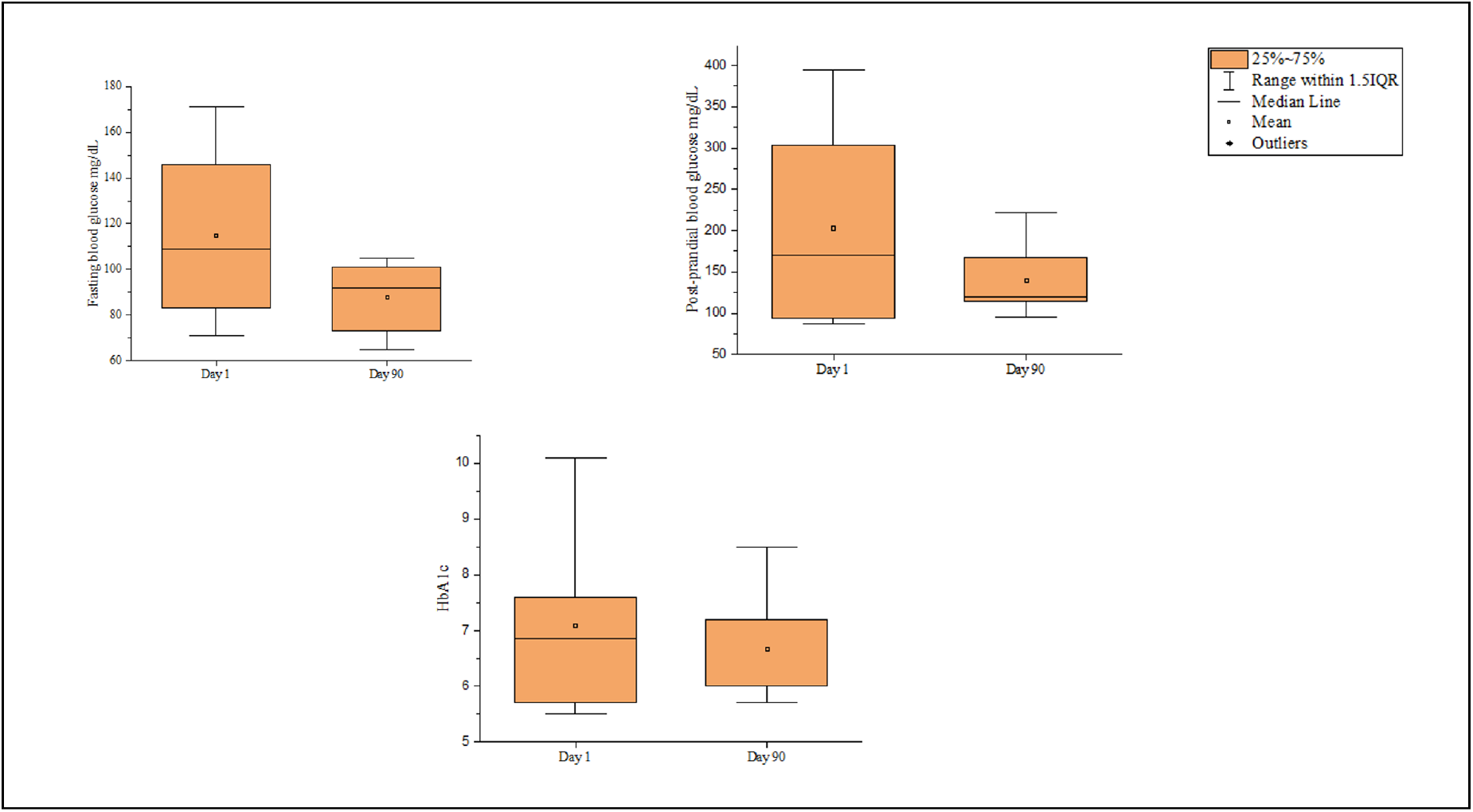
Values of fasting, post-prandial blood glucose, and HbA1c pre and post consumption of AFO-202 beta-glucan

MRI was used to calculate the Parkinsonism Index (2.0); in one patient, the MRPI (P/M) × (MCP/SCP) decreased from 9.3 at baseline to 9.1 post intervention, and the MRPI 2.0 – MRPI × (V3/FH) remained the same (0.7) at baseline and after 90 days (Figure 5). MRPI did not show any improvement in the other patients.

**Figure 5:**
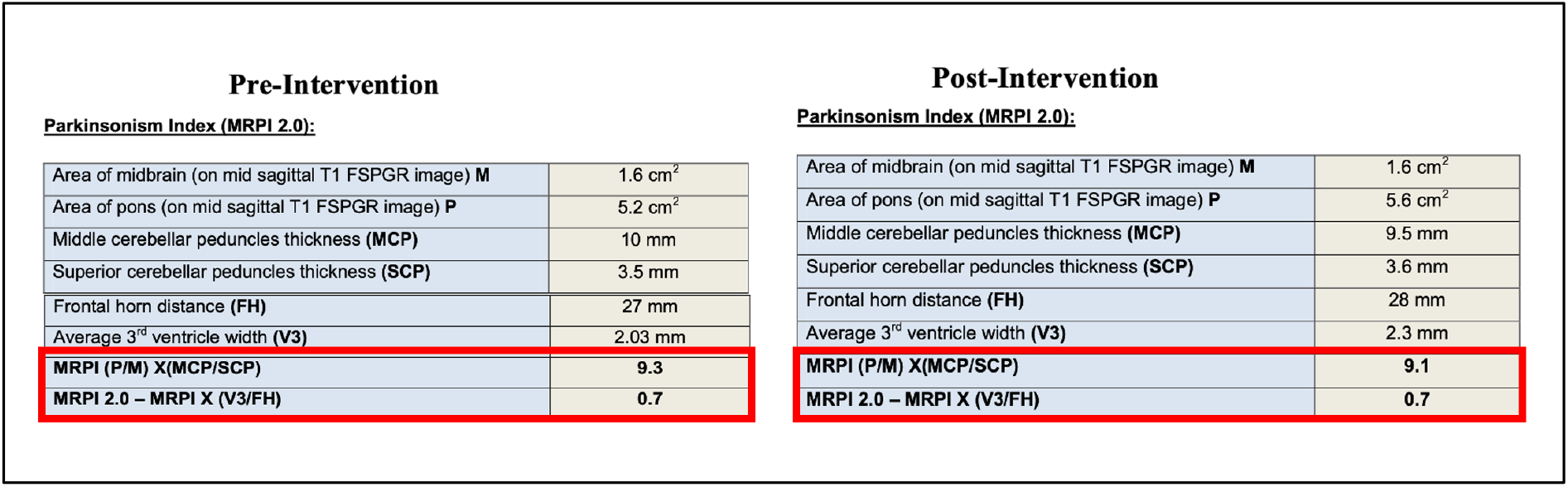
Magnetic Resonance Imaging Parkinsonism Index (2.0) (MRPI), which improved in one patient

There were no serious adverse effects in any of the study subjects.

## Discussion

PD is a multifactorial disease, and its pathogenesis is intricately influenced by environmental variables, chemical exposure (insecticides and pesticides), xenobiotic toxins, genetic predisposition, altered dopamine metabolism, mitochondrial dysfunction, oxidative stress, neuroinflammation, and aging. While key motor characteristics of PD include rigidity, slowness of movement, resting tremors, and abnormal gait, the majority of PD patients notice non-motor symptoms (NMS), such as gastrointestinal (GI) symptoms of dry mouth, constipation, and defaecatory dysfunction, decades before motor symptoms manifest [9]. Thus, gut dysbiosis plays a central role in the pathogenesis of PD. Because the gut microbiota and its metabolites control neuroinflammation, barrier function, and neurotransmitter activity, it has been hypothesised that these factors contribute to the aetiology of PD. The microbiota-gut-brain axis may offer a route for the transmission of misfolded α-synuclein because of the bidirectional contact between the enteric nervous system and central nervous system (CNS) in humans [10]. α-Synuclein misfolding and aberrant aggregation are caused by changes in the gastrointestinal microbiome (GM) in the intestine, especially an increase in pathogenic bacteria such as Enterobacteriaceae that produce curli proteins and abnormal amyloids. The vagus nerve carries abnormal α-synuclein into the central nervous system (CNS) because it is not removed by physiological mechanisms. In addition to causing the production of eosinophilic Lewis Bodies in the cytoplasm and mitochondrial malfunction in dopaminergic (DA) neurones, abnormal amounts of α-synuclein aggregating in the substantia nigra pars compacta stimulate an inflammatory response in the microglia. An increase in oxidative stress (OS) caused by these pathogenic modifications leads to the death of nerve cells, which is a PD-associated trait. This increase in OS amplifies and further oxidises the aberrant aggregation of α-synuclein, creating a positive feedback loop [10,11].

Therefore, disease-modifying approaches are necessary to address the GI symptoms of constipation as well as to regulate α-synuclein. In the current study, there was a significant improvement in the constipation severity scale, with patients reporting subjective improvement in quality of life due to the same after AFO-202 beta-glucan consumption. The primary outcome metric in Parkinson’s clinical trials was the UPDRS [12]. A change of eight points in the UDPRS total score represented a clinically significant improvement in early PD after six months of treatment, while a change of five points in the UPDRS motor portion was shown to be the most acceptable cutoff score. However, this improvement was observed after six months of treatment. In the current study, although there was only a modest improvement in the UPDRS score, it was observed over a span of three months, warranting longer duration studies of AFO-202 beta-glucan in PD.

Patients with Parkinson’s disease have been found to have higher levels of serum creatine kinase [13]. Patients with PD may react negatively to physical exercise, causing CK to leak from skeletal muscles. Elevated serum CK levels may also be caused by neural mechanisms regulated by hypothalamic dopamine and the autonomic nervous system. In the present study, serum CK levels decreased post intervention.

PD and type 2 diabetes mellitus (T2DM) are co-occurring conditions that can have adverse effects on cognition and non-motor and motor symptoms [14]. Patients with T2DM experience significant gait impairment and mild cognitive impairment more quickly than those without. Therefore, addressing the diabetic component of PD has been suggested as a potential approach to slowing disease progression. In the current study, the reduction in fasting and postprandial blood glucose levels after AFO-202-glucan consumption adds to the value of this beta-glucan as a potential DMT adjuvant in the treatment of PD.

The limitations of the study are the small sample size and short duration (three months. However, it has been reported [15] that because PD is a slowly developing condition, disease-modifying trials require lengthy follow-up intervals, and enrolling large populations with longer durations, along with the disease’s heterogeneity, raises the cost of clinical trials.

Furthermore, specific gut microbiome sequencing of these patients in future studies will be critical to decipher the underlying complex pathogenic mechanisms, in addition to designing novel approaches to address PD in a prophylactic as well as therapeutic manner. Even in the patients who were included in the study, since PD has been present for a long time, the already-established abnormal amyloid deposition in the brain and how far the present therapeutic approaches will help remain unknown. The role of microglia is important in this context, as abnormal α-synuclein inhibits microglial phagocytosis by binding to the Fc gamma receptor IIB (FcRIIB) on their surface, which may make it more difficult to clear aggregated species or other parenchymal wastes. In addition, the fibrillar α-synuclein stimulates the NF-B pathway in microglia, which is essential for inflammatory microglial responses [16]. In an earlier study, beta-glucan has been shown to prevent the high-fat and fibre-deficient diet (HFFD)-induced activation of the microglia, their engulfment of synaptic puncta, and an increase in the mRNA expression of proinflammatory cytokines (TNF-, IL-1, and IL-6) in the hippocampus. Another mechanism involves GABAergic interneurons, as α-synuclein accumulation and beta-III tubulin protein complex regulate the release of synaptic vesicles and lower GABAergic inhibitory transmission, which leads to neuronal dysfunction [17]. Therefore, the positive effects of beta-glucan in patients with established PD in the present study can also be attributed to the proper activation of microglia, apart from the regulation of GABAergic inhibitory transmission [18], as well as the action through the serotonergic system [19]. In patients with young-onset PD and those where a specific causal relationship between antibiotics and PD due to gut microbial resistance is suspected [20], this beta-glucan based approach can also be prophylactic, addressing the pathogenic mechanisms in a continuous systemic and local manner, which needs further research for validation.

## Conclusion

This study demonstrated the safety and beneficial disease-modifying potential of the *A. pullulans* AFO-202 strain produced beta-glucans in patients with PD over a short span of 90 days. Improvements in the UPDRS, CSS, and MRI were clinically significant, and a larger multicentre study is recommended to validate the novel Nichi BRITE beta-glucans as an adjuvant in the management of PD. In such larger studies, evaluation of gut microbiome in response to NICHI BRITE beta-glucans is also needed to ascertain their earlier proven beneficial gut microbiome reconstitution for application in neurological illnesses such as synculeinopathies as well Lewy-body dementia.

## Data Availability

All data produced in the present work are contained in the manuscript

## Acknowledgments

The authors would like to

1. Mr. Yasushi Onaka and Mr. Masato Onaka, of Sophy Inc. for technical clarifications,
2. Ms. Eiko Amemiya of II Dept. of Surgery, University of Yamanashi for secretarial assistance.
3. Dr. Vellaichamy Kannan, Neurologist, for neurological evaluations and patient recruitment.
4. Dr. Malar Vetrievel, BeWell Hospitals for clinical trial coordination.
5. Dr. Suryakumar, Radiologist, Anderson Diagnostics for MRPI Index evaluation.
6. Dr. Ragaroobine, Mr. Mathaiyan Rajmohan of NCRM, Dr. Subbiah Sriraam, Ms.Aditi Ramesh and staff of Aurous HealthCare for their assistance with the regulatory procedures, documentation, logistics and data collection of the study.
7. Mr. Yoshio Morozumi and Ms. Yoshiko Amikura of GN Corporation, Japan for liaising between the institutes
8. Loyola-ICAM College of Engineering and Technology (LICET) for their overall support to our research work.

## References

1. Stoker TB, Barker RA. Recent developments in the treatment of Parkinson’s Disease. F1000Res. 2020 Jul 31;9:F1000 Faculty Rev-862.

2. Kalia LV, Lang AE: Parkinson’s disease. Lancet. 2015;386(9996):896–912. 10.1016/S0140-6736(14)61393-3

3. Dickson DW: Parkinson’s disease and parkinsonism: Neuropathology. Cold Spring Harb Perspect Med.2012;2(8):a009258. 10.1101/cshperspect.a009258

4. Fields CR, Bengoa-Vergniory N, Wade-Martins R: Targeting Alpha-Synuclein as a Therapy for Parkinson’s Disease. Front Mol Neurosci. 2019;12:299. 10.3389/fnmol.2019.00299

5. Araki K, Yagi N, Aoyama K, Choong CJ, Hayakawa H, Fujimura H, Nagai Y, Goto Y, Mochizuki H. Parkinson’s disease is a type of amyloidosis featuring accumulation of amyloid fibrils of α-synuclein. Proc Natl Acad Sci U S A. 2019 Sep 3;116(36):17963–17969.

6. Raghavan K, Dedeepiya VD, Kandaswamy RS, Balamurugan M, Ikewaki N, Sonoda T, Kurosawa G, Iwasaki M, Preethy S, Abraham SJ. Improvement of sleep and melatonin in children with autism spectrum disorder after β-1,3/1,6-glucan consumption: An open-label prospective pilot clinical study. Brain Behav. 2022 Sep;12(9):e2750

7. Raghavan K, Dedeepiya VD, Ikewaki N, Sonoda T, Iwasaki M, Preethy S, Abraham SJ. Improvement of behavioural pattern and alpha-synuclein levels in autism spectrum disorder after consumption of a beta-glucan food supplement in a randomised, parallel-group pilot clinical study. BMJ Neurol Open. 2022 Jan 18;4(1):e000203.

8. Raghavan K, Dedeepiya VD, Yamamoto N, Ikewaki N, Sonoda T, Iwasaki M, Kandaswamy RS, Senthilkumar R, Preethy S, Abraham SJK. Benefits of Gut Microbiota Reconstitution by Beta 1,3-1,6 Glucans in Subjects with Autism Spectrum Disorder and Other Neurodegenerative Diseases. J Alzheimers Dis. 2022 Sep 6. doi: 10.3233/JAD-220388.

9. Pavan S, Prabhu AN, Prasad Gorthi S, Das B, Mutreja A, Shetty V, Ramamurthy T, Ballal M. Exploring the multifactorial aspects of Gut Microbiome in Parkinson’s Disease. Folia Microbiol (Praha). 2022 Oct;67(5):693–706.

10. Lei Q, Wu T, Wu J, Hu X, Guan Y, Wang Y, Yan J, Shi G. Roles of α-synuclein in gastrointestinal microbiome dysbiosis-related Parkinson’s disease progression (Review). Mol Med Rep. 2021 Oct;24(4):734.

11. Wang Q, Luo Y, Ray Chaudhuri K, Reynolds R, Tan EK, Pettersson S. The role of gut dysbiosis in Parkinson’s disease: mechanistic insights and therapeutic options. Brain. 2021 Oct 22;144(9):2571–2593.

12. Schrag A, Sampaio C, Counsell N, Poewe W. Minimal clinically important change on the unified Parkinson’s disease rating scale. Mov Disord. 2006 Aug;21(8):1200–7.

13. Takubo H, Shimoda-Matsubayashi S, Mizuno Y. Serum creatine kinase is elevated in patients with Parkinson’s disease: a case controlled study. Parkinsonism Relat Disord. 2003 Apr;9 Suppl 1:S43–6.

14. Mischley LK, Lau RC, Bennett RD. Role of Diet and Nutritional Supplements in Parkinson’s Disease Progression. Oxid Med Cell Longev. 2017;2017:6405278.

15. Athauda D, Evans J, Wernick A, Virdi G, Choi ML, Lawton M, Vijiaratnam N, Girges C, Ben-Shlomo Y, Ismail K, Morris H, Grosset D, Foltynie T, Gandhi S. The Impact of Type 2 Diabetes in Parkinson’s Disease. Mov Disord. 2022 Aug;37(8):1612–1623.

16. Kam TI, Hinkle JT, Dawson TM, Dawson VL. Microglia and astrocyte dysfunction in parkinson’s disease. Neurobiol Dis. 2020 Oct;144:105028. doi: 10.1016/j.nbd.2020.

17. Shi H, Yu Y, Lin D, Zheng P, Zhang P, Hu M, Wang Q, Pan W, Yang X, Hu T, Li Q, Tang R, Zhou F, Zheng K, Huang XF. β-glucan attenuates cognitive impairment via the gut-brain axis in diet-induced obese mice. Microbiome. 2020 Oct 2;8(1):143.

18. Ito H, Nakayama K, Jin C, Suzuki Y, Yazawa I. α-Synuclein accumulation reduces GABAergic inhibitory transmission in a model of multiple system atrophy. Biochem Biophys Res Commun. 2012 Nov 23;428(3):348–53.

19. Dutta SD, Patel DK, Ganguly K, Lim KT. Effects of GABA/β-glucan supplements on melatonin and serotonin content extracted from natural resources. PLoS One. 2021 Mar 5;16(3):e0247890.

20. Mertsalmi TH, Pekkonen E, Scheperjans F. Antibiotic exposure and risk of Parkinson’s disease in Finland: A nationwide case-control study. Mov Disord. 2020 Mar;35(3):431–442.

